# The Relationship between Alcohol- and Sleep-related Traits: Results from Polygenic Risk Score Analyses and Mendelian Randomization Studies

**DOI:** 10.1101/2023.02.19.22282910

**Authors:** Subhajit Chakravorty, Rachel L. Kember, Diego R. Mazzotti, Hassan S. Dashti, Sylvanus Toikumo, Philip R. Gehrman, Henry R. Kranzler

## Abstract

**Background:** Epidemiologic studies have shown an association between sleep abnormalities and alcohol-related traits. Recent genome-wide association studies (GWAS) have identified genetic variants associated with sleep-related traits, including insomnia and sleep duration, and with alcohol-related phenotypes, including alcohol use disorder (AUD) and level of alcohol consumption.

**Objectives:** We investigated whether genetic risk for insomnia and sleep duration abnormalities are associated with AUD and alcohol consumption. We also evaluated the causal relationships between sleep- and alcohol-related traits.

**Methods:** Individual level phenotype and genotype data from the Million Veteran Program was used. Polygenic risk scores (PRS) were computed using summary statistics from two recent discovery GWAS of insomnia (N=453,379 European-ancestry (EA) individuals) and sleep duration (N=446,118 EAs) and tested for association with lifetime AUD diagnosis (cases, N=34,658 EAs) and past-year Alcohol Use Disorders Identification Test-Consumption scale scores (AUDIT-C, N=200,680 EAs). Bi-directional two-sample Mendelian Randomization (MR) analyses assessed causal associations between the two sleep traits and the two alcohol-related traits.

**Results:** Insomnia PRS was positively associated with AUD at 2/9 PRS thresholds, with p<0.01 being the most significant (OR = 1.02, p = 3.48 × 10^−5^). Conversely, insomnia PRS was negatively associated with AUDIT-C at 6/9 PRS thresholds (most significant threshold being p=0.001 (β = - 0.02, p = 5.6 × 10^−8^). Sleep duration PRS was not associated with AUD, but was positively associated with AUDIT-C at 2/9 PRS thresholds, with the most significant threshold being p = 1 × 10^−6^ (β = 0.01, p = 0.0009). MR analyses supported a significant positive causal effect of insomnia on AUD (14 SNPs; beta = 104.14; SE = 16.19; p = 2.22 × 10^−5^), although with significant heterogeneity. MR analyses also provided nominal evidence of a causal effect of AUD on insomnia (10 SNPs; beta = 0.01; SE = 0.007; p = 0.01). Finally, MR analyses showed that decreased sleep duration had a causal effect on the risk of AUD (27 SNPs; beta = -63.05; SE = 3.54; p = 4.55 × 10^−16^) and was robust to sensitivity analyses.

**Conclusion:** The genetic risk for insomnia shows pleiotropy with AUD, and sleep continuity abnormalities have a causal influence on the development of AUD.

## Introduction

Alcohol is a psychoactive substance that is consumed by nearly two-thirds of adults in the United States (US). ^1^ Among these individuals, almost a third (29.9%) indulge in heavy drinking, defined as the consumption of ≥14 standard drinks a week or >4 drinks on any day in men and ≥ 7 drinks a week or >3 drinks on any day in women.^2^ Heavy drinking is a potentially modifiable risk factor for traumatic injuries and medical, psychiatric, and sleep-related disorders.^3-5^ Insomnia and short (insufficient) sleep duration are the most common sleep-related disorders that accompany heavy alcohol consumption and alcohol use disorder (AUD). Insomnia is a disorder of sleep continuity that consists of difficulty falling asleep or staying asleep, or early morning awakening that leads to impaired functioning. Insomnia has a bi-directional causal relationship with alcohol consumption^6^ and is also a risk factor for the development of AUD,^7^ but it is unclear whether AUD is causal in the development of insomnia.

Sleep duration, another aspect of sleep health, is the total duration of sleep obtained, either during nocturnal sleep or across a 24-hour period^8^ with a range of 7-9 hours recommended to support optimal health in adults.^9^ Abnormalities of sleep duration, especially short sleep duration (≤6 hours a night), are associated in cross-sectional studies with an increased risk of adverse health consequences such as mortality, suicide, physical injuries, cardio-metabolic and psychiatric comorbidities,^10^ and heavy drinking and AUD.^11-15^ It is unclear whether short sleep duration is a cause or a consequence of heavy drinking or AUD.

Genetic factors influence variation in both alcohol- and sleep-related traits.^16^ Recent estimates of the SNP-based heritability (i.e., that attributable to common genetic variation) among European-ancestry (EA) individuals is 7-11% for alcohol consumption (based on the Alcohol Use Disorders Identification Test-Consumption (AUDIT-C) measure),^17,18^ 6-16% for AUD,^17,19^ 7-17% for insomnia,^20,21^ and 10% for sleep duration.^22^ A polygenic risk score (PRS), an individual’s estimated genetic predisposition for a given trait, is computed as the sum of the alleles associated with the trait, weighted by the risk allele effect sizes.^23^ PRS have been used to validate genetic links to disease, dissect pleiotropic associations across traits, and provide a quantitative measure of an aggregated genetic burden for illness in an individual. Here, we evaluated the associations between insomnia or sleep duration-related PRS and two alcohol-related phenotypes (AUD and alcohol consumption).

The availability of large biobanks such as the United Kingdom Biobank (UKB) and the Million Veteran Program (MVP), which include a range of phenotypic data, has allowed researchers to conduct genome-wide association studies (GWAS) for traits like insomnia, sleep duration, AUD, and alcohol consumption.^20-22^ In addition to serving as discovery samples for the calculation of PRS, summary statistics from these GWASs can be used to conduct Mendelian Randomization (MR) studies to understand causal relationships between traits. MR studies can provide evidence supportive of causal relations because genetic variants are not substantially modifiable and are free from reverse confounding.^24,25^ An MR study uses genetic variation identified in GWAS to create exposure and outcome instruments that determine the causal effect of the exposure (e.g., insomnia) on an outcome (e.g., AUD).^25,26^

A preliminary study of the relationship between insomnia and AUD using MR analysis showed a significant causal effect of insomnia on the development of AUD in a GWAS study of 46,568 individuals with the disorder.^27^ We repeated this analysis using data from a GWAS of AUD comprising 202,004 individuals, to evaluate the bidirectional relationship between insomnia and AUD. We also assessed bidirectional relationships between insomnia and alcohol consumption using data from a GWAS of alcohol consumption involving 200,680 individuals and between habitual reported sleep duration and both AUD and alcohol consumption. We evaluated AUD and AUDIT-C as separate alcohol-related traits as they have shown genetic independence in their correlation with non-alcohol-related traits and the phenotypes associated with their traits’ PRS^17^ and thus may have different associations with sleep-related phenotypes. Findings from these analyses improve our understanding of the relations between sleep- and alcohol-related disorders and can help to guide the development of individualized care at this interface.^28^

## Methods

### Overview

We used GWAS summary statistics from the UKB and individual-level data from MVP to construct polygenic risk scores for insomnia and sleep duration and test their association with alcohol-related phenotypes.^23^ We used MVP GWAS summary statistics for alcohol-related phenotypes and UKB summary statistics for insomnia and sleep duration to test their causal relations.

#### Standard protocols and informed consent

We used summary level data from the UKB, a large biobank study investigating the contributions of genetic predisposition and environmental exposure to the development of disease. All UKB subjects provided written informed consent to participate and make their de-identified data publicly available.^29^ The MVP is an observational cohort study and biobank supported by the U.S. Department of Veterans Affairs (VA).^30^ Phenotypic data were collected from MVP subjects using questionnaires, the VA electronic health record (EHR), and a blood sample for genetic analysis. The MVP provided access to patient-level AUDIT-C scores and AUD diagnoses, GWAS summary statistics, and approved the data analysis. The MVP study followed all relevant ethical regulations for research with human subjects and obtained informed consent from all participants.

### Data source

#### Insomnia GWAS

We obtained summary statistics from the published GWAS of insomnia in EA participants in the UKB (N=453,379).^21^ This study assessed insomnia symptoms using the question (data field 1200), “Do you have trouble falling asleep at night or do you wake up in the middle of the night?” The responses were primarily evaluated as a binary case-control variable, with cases reporting insomnia symptoms as “usually” or “sometimes” and controls endorsing “never/rarely.”

#### Sleep duration GWAS

We obtained the GWAS summary statistics for sleep duration in EA participants from the UKB (N=446,118). The study assessed sleep duration using the question (data field 1160), “About how many hours sleep do you get in every 24 hours?” The response was recorded as a whole integer within a range of 1 to 23, but responses <3 and >18 hours were excluded from the GWAS analysis.^22^

#### Alcohol-related GWAS

We used the GWAS summary statistics for AUDIT-C score and AUD in EA participants from the MVP (AUDIT-C [N= 200,680]; AUD [N = 202,004]).^17^

#### Alcohol phenotypes in the Million Veteran Program (MVP)

We used the individual-level AUDIT-C scores^31^ and AUD diagnostic codes from the Department of Veterans Affairs (VA) EHR. The AUDIT-C consists of the first three items of the Alcohol Use Disorders Identification Test and measures typical quantity (question 1) and frequency (question 2) of drinking, and frequency of heavy drinking (question 3). The AUDIT-C is a mandatory annual assessment for all veterans seen in primary care within the VA health system. Alcohol-related disorders were identified using International Classification of Diseases (ICD), 9^th^ revision (ICD-9) codes 303.X (dependence) and 305–305.03 (abuse) and ICD, 10^th^ revision (ICD-10) codes F10.1 (abuse) and F10.2 (dependence). EA participants with at least one inpatient or two outpatient alcohol-related ICD-9/10 codes (N = 34,658) from 2000 to 2018 were assigned a diagnosis of AUD.^17^

#### Genetic Correlations

We used the cross-trait Linkage Disequilibrium (LD) score regression to calculate genome-wide genetic correlations (r_g_) between insomnia, sleep duration, and alcohol use phenotypes, using precomputed LD scores for European individuals from the 1000 Genomes Project.^32^

#### Polygenic risk scores (PRS)

We used the clumping and thresholding method^23^ to create the PRS for insomnia and sleep duration in EA individuals (N=209,020). The PRS was calculated using the dosage information for each variant under an additive model as the sum of all alleles carried, weighted by the effect size of the allele in the GWAS. We performed p-value informed clumping using the 1000 Genomes EA participants as the LD background, with r^2^ = 0.1 and a distance threshold of 250 kb. The PRS was computed for nine p-value thresholds (p ≤ 0.000001, 0.00001, 0.0001, 0.001, 0.01, 0.05, 0.1, 0.5, and 1) and standardized the PRS with a mean = 0, and SD = 1. Linear regression (for AUDIT-C score as the outcome) and binary logistic regression (for AUD cases and controls) evaluated their association with PRS as the predictor after adjusting for covariates that included age, sex, and the first five principal components of their genetic ancestry (PCs) (Figure 1). A Bonferroni-corrected p-value (0.05/9=0.0056) was used to account for multiple testing.

**Figure 1.**
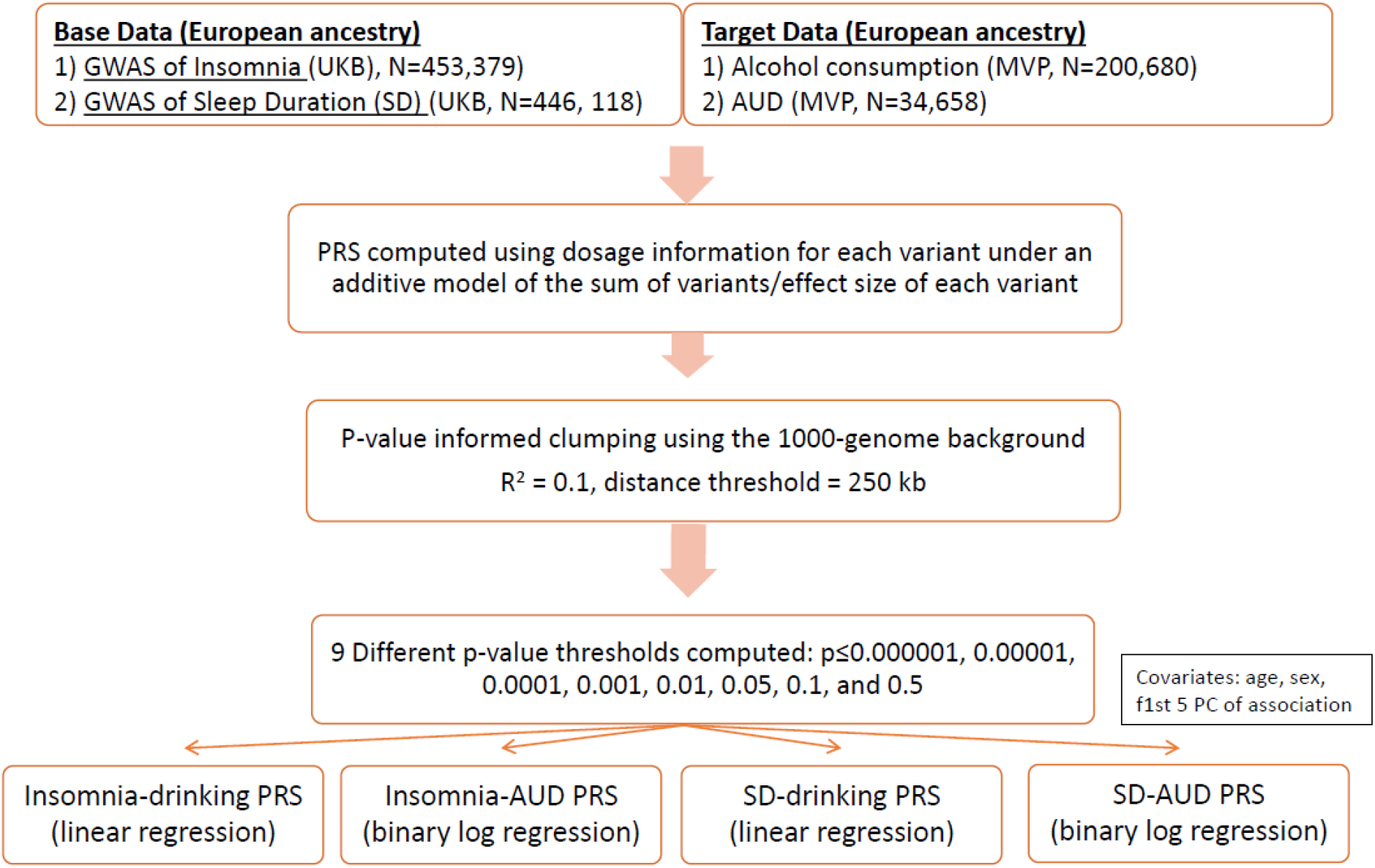
PRS methodology in the current study **Footnotes:** GWAS = genome-wide association study; UKB = United Kingdom Biobank; MVP = Million Veteran Program; PRS = Polygenic Risk Score; AUD = Alcohol Use Disorder; PC = Principal Components;

#### Mendelian randomization analyses (MR)

We harmonized the GWAS summary statistics for insomnia and alcohol-related phenotypes before conducting genetic correlation analyses, bidirectional MR analyses, and sensitivity analyses. The primary MR method was random-effects inverse-variance weighted (IVW) regression for balanced horizontal pleiotropy and mode-based estimator for majority horizontal pleiotropy^25,33^ with sleep (duration or insomnia) and alcohol-related variable (AUDIT-C score or AUD diagnosis) alternately used as exposure or outcome. We employed a stringent clumping threshold of r^2^ = 0.001. For dichotomous phenotypes (insomnia and AUD), a one-unit increase in the genetic instrument reflects a doubling in the odds of the exposure trait.^34^ MR provides strong evidence for causality provided the following assumptions are met for the genetic instrument: (1) it is strongly associated with the exposure, (2) it is not associated with confounders, and (3) there is no horizontal pleiotropy, i.e., it only affects the outcome through its effect on the exposure.^24^ We performed sensitivity analyses using more robust genetic instruments for insomnia to address the first assumption. The use of randomly distributed alleles as the instrumental variables and the adjustment for population stratification in the GWAS satisfied the second assumption. We used multiple models robust to various forms of pleiotropy to evaluate the third assumption. The MR analyses included two studies of the causal effect of insomnia on alcohol consumption and AUD and two studies of the causal effect of alcohol consumption and AUD on insomnia (Figure 2). Thus, the four insomnia-related MR analyses yielded a Bonferroni-adjusted alpha threshold of 0.05/4 = 0.0125. Similarly, the four separate sleep duration-related MR analyses used a Bonferroni-adjusted alpha threshold of 0.05/4 = 0.0125. P-values lower than the corrected alpha thresholds represent significant evidence for causal effects, and p<0.05 provides nominal evidence for a causal effect. We used Cochrane’s Q-statistic to assess SNP effect heterogeneity^35^ and single-SNP analysis and the leave-one-out IVW analyses to evaluate the disproportionate effects of single SNPs in the models.^25^ The F-scores for the sleep and alcohol-related instruments ranged from 40.06 – 90.71. Wherever horizontal pleiotropic effects were suspected based on the scatter plot results, median or mode-based estimators were applied, as applicable. The median-based approach provides an unbiased estimate in the presence of unbalanced horizontal pleiotropy, and the causal estimate from the mode-based estimator is unbiased if the SNPs contributing to the largest cluster are valid instruments. ^25^

**Figure 2.**
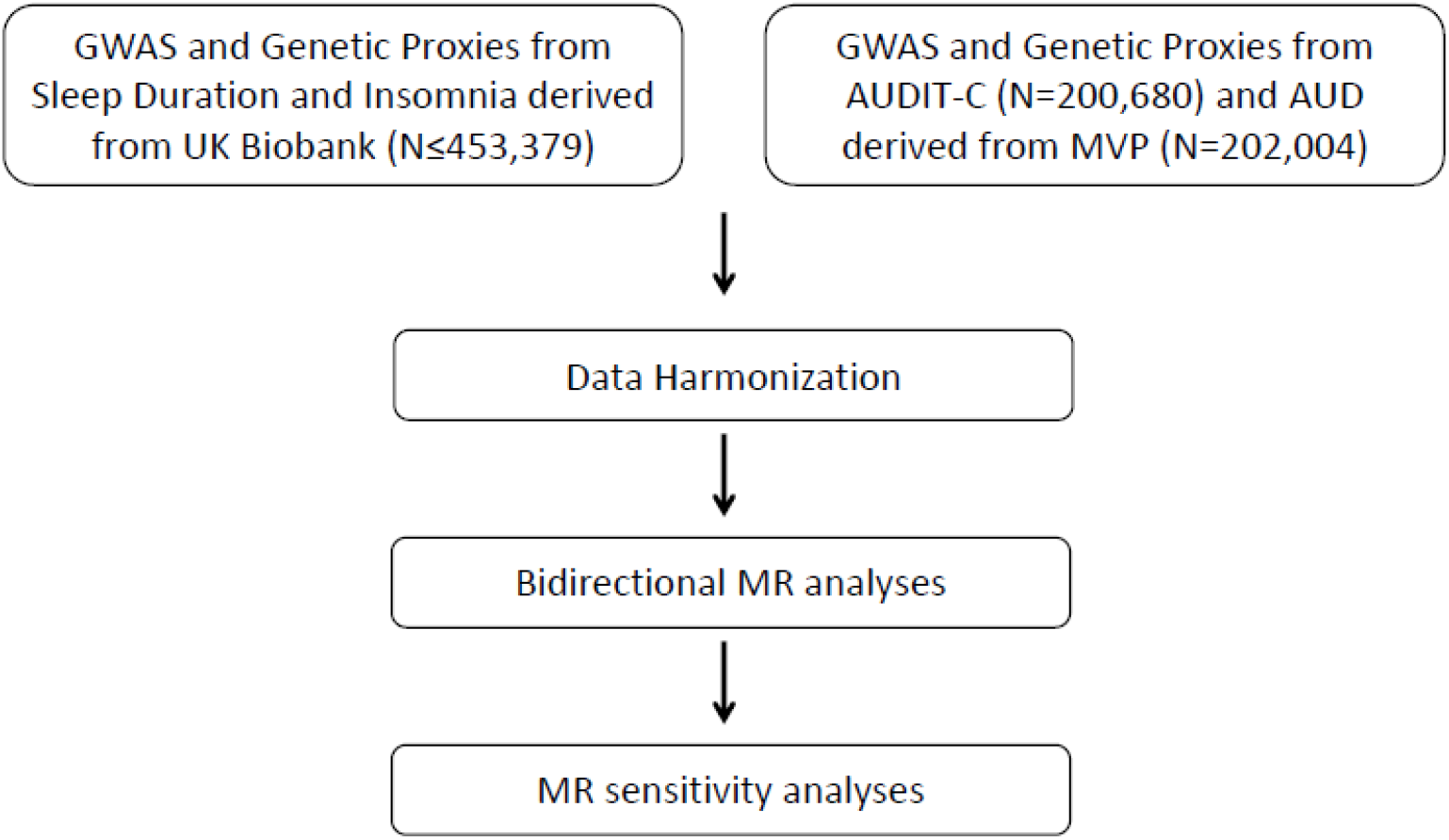
Mendelian Randomization analysis pipeline in the current study **Footnote:** GWAS = gene-wide association studies; AUDIT-C = Alcohol Use Disorder Identification Test – Consumption (a measure for drinking); AUD = Alcohol Use Disorder (“alcoholism”); MVP = Million Veteran Program; PC = Principal Components; MR = Mendelian Randomization study.

#### Statistical Software

The genetic correlations were analyzed using LDSC v1.01,^32^ PRS analysis was conducted using PLINK version 2,^36^ and MR analysis was performed in R version 4.1.2^37^ using the TwoSampleMR package version 0.5.6.^25^

## Results

### Genetic Correlations

Insomnia was positively genetically correlated with AUD (r_g_ = 0.17, p = 3.00 × 10^−8^), but negatively correlated with AUDIT-C (r_g_ = -0.15, p = 1.27 × 10^−5^). Sleep duration was positively correlated with AUDIT-C (r_g_ = 0.12, p = 1.59 × 10^−4^) but not AUD (r_g_ = 0.05, p = 9.99 × 10^− 2^; supplementary table 1.

### Insomnia

#### Insomnia PRS is associated with alcohol consumption and AUD

At two of the PRS thresholds there was a significant positive association of insomnia PRS with AUD, with the most significant being p<0.01 [Odds Ratio (OR) = 1.02, p = 3.48 × 10^−5^, Figure 3a and Supplementary Table 2]. In contrast, insomnia PRS was significantly associated with lower AUDIT-C scores for six of the nine thresholds, the most significant of which was p=0.001 (Beta = -0.02, p = 5.6 × 10^−8^; Figure 3b and Supplementary Table 2). A secondary analysis showed no relationship between the mean insomnia PRS score and the individual AUDIT-C scores from 0-12 (Supplementary Figure 1).

**Figure 3:**
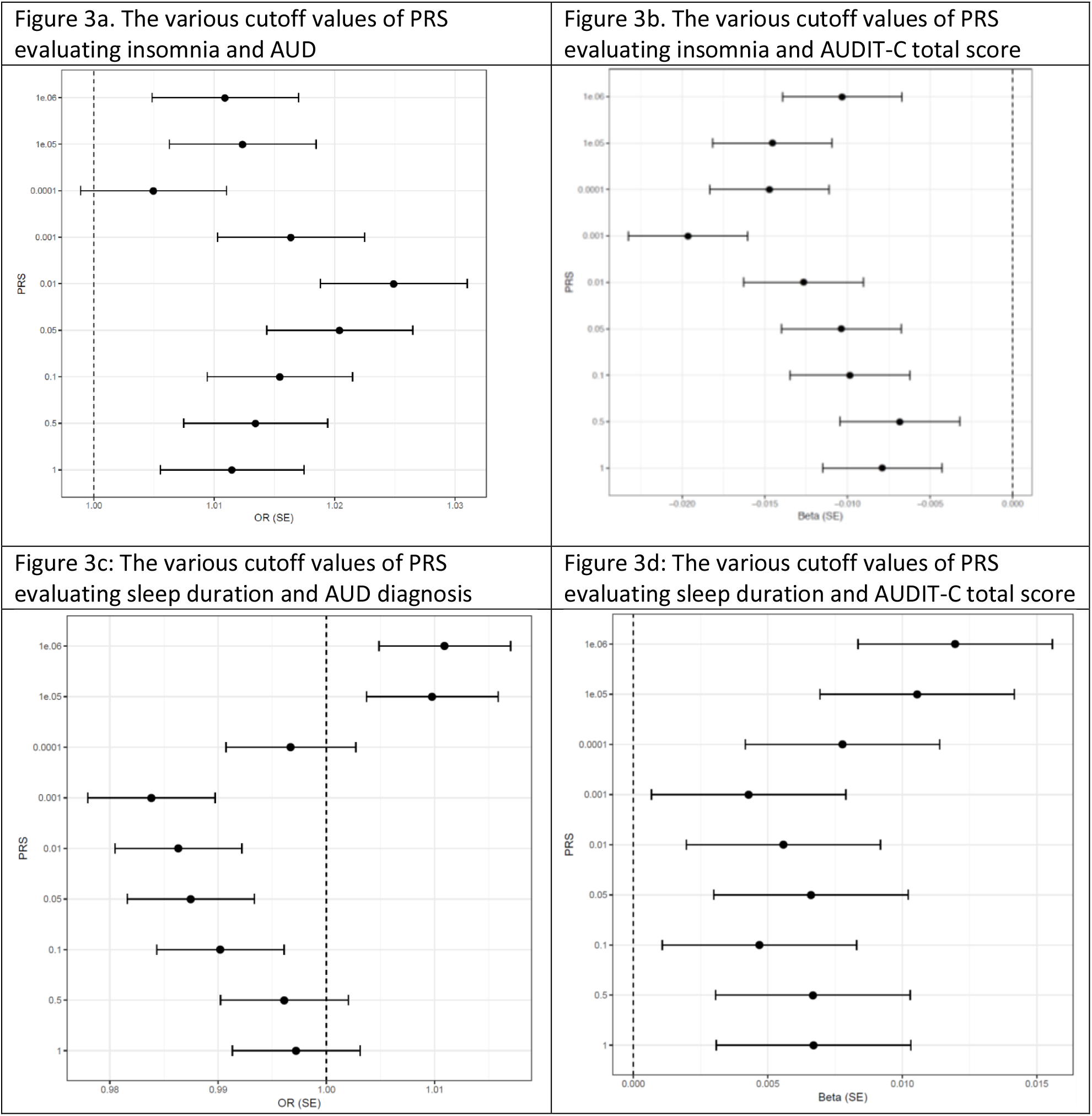
Polygenic Risk Score Analysis of sleep- and alcohol-related variables **Footnote:** PRS = polygenic risk score; AUD = Alcohol Use Disorder; AUDIT-C = Alcohol Use Disorder Identification Test – Consumption; Beta = coefficient of regression; OR = odds ratio; SE = standard error.

#### Mendelian randomization analyses support the causal effects of insomnia on AUD

The scatter plot showed heterogeneity and the resultant mode-based estimator showed a significant positive causal effect of insomnia on AUD (14 SNPs; beta = 104.14; SE = 16.19; p = 2.22 × 10^−5^, Figure 4, Supplementary Table 3, and Supplementary Figure 2). There was a significant weighted mode estimator in the sensitivity analyses for insomnia as an exposure (beta = -82.62, SE = 5.50, p = 1.38 × 10^−9^, Supplemental Figure 2). The leave-one-out analysis did not support excluding any SNPs from the analysis (Supplementary Table 3, Supplementary Figure 3). All 14 single-SNP analyses showed significant correlations, with p-values that ranged between 2.42 × 10^−8^ to 1.08 × 10^−16^. Of these, 7 SNPs were positively correlated, and 7 SNPs were negatively correlated. There was heterogeneity (Cochrane’s Q test p<0.0001) but no evidence of horizontal pleiotropy (p = 0.69).

**Figure 4.**
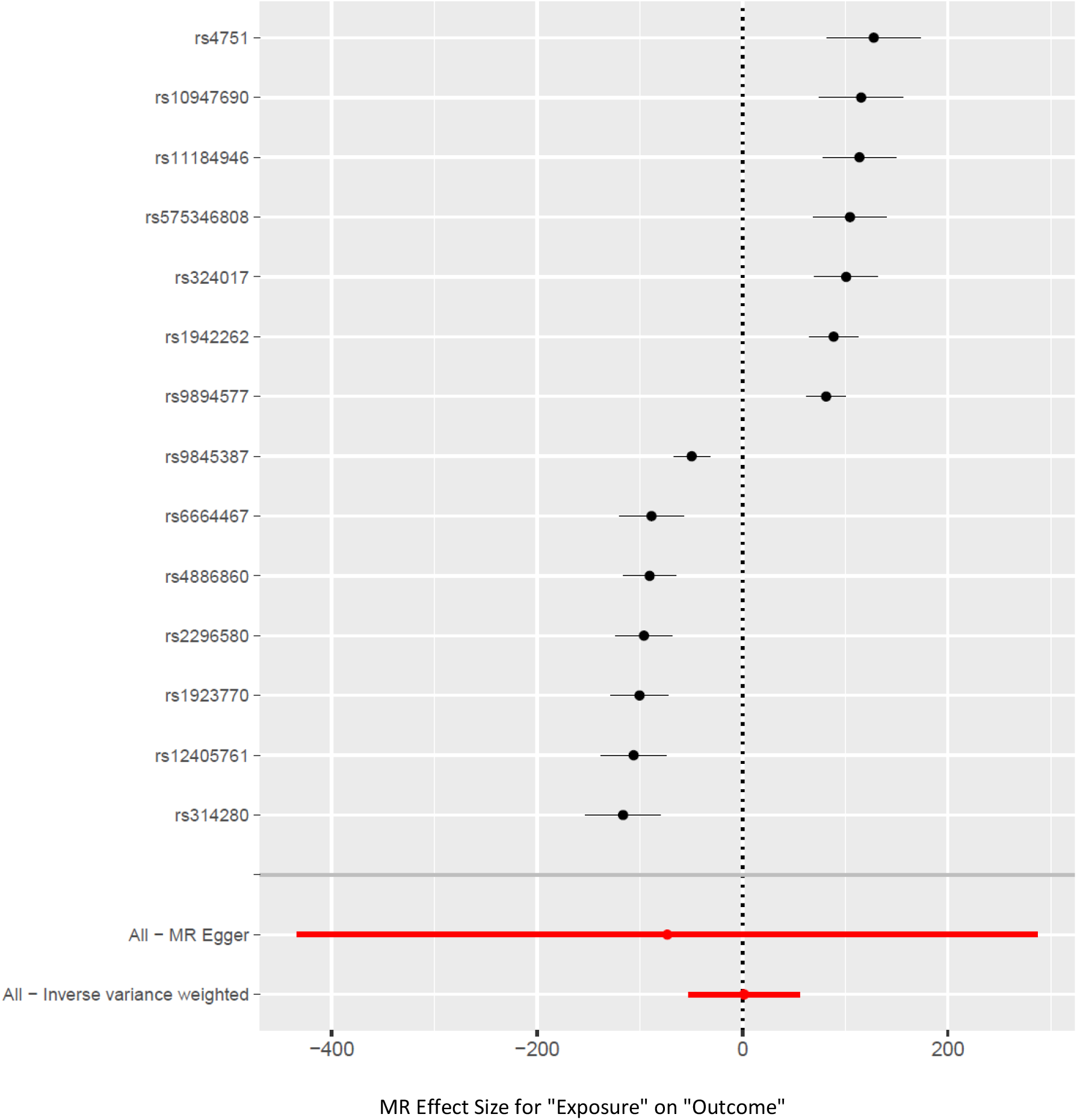
Forest Plot of the causal effect of Insomnia (exposure) on Alcohol Use Disorder (AUD, outcome)

#### Other findings

The weighted median estimator showed a nominally positive causal effect of AUD on insomnia (10 SNPs; beta = 0.01; SE = 0.007; p = 0.01; Supplementary Table 4 and Supplementary Figures 4 and 5) although there was heterogeneity in the data (Cochrane’s Q test p = 6.24 × 10^−8^) but no horizontal pleiotropic effects. Sensitivity analyses for AUD showed a significant weighted mode estimator (beta = 0.02, SE = 0.008, p = 0.02).

**Figure 5.**
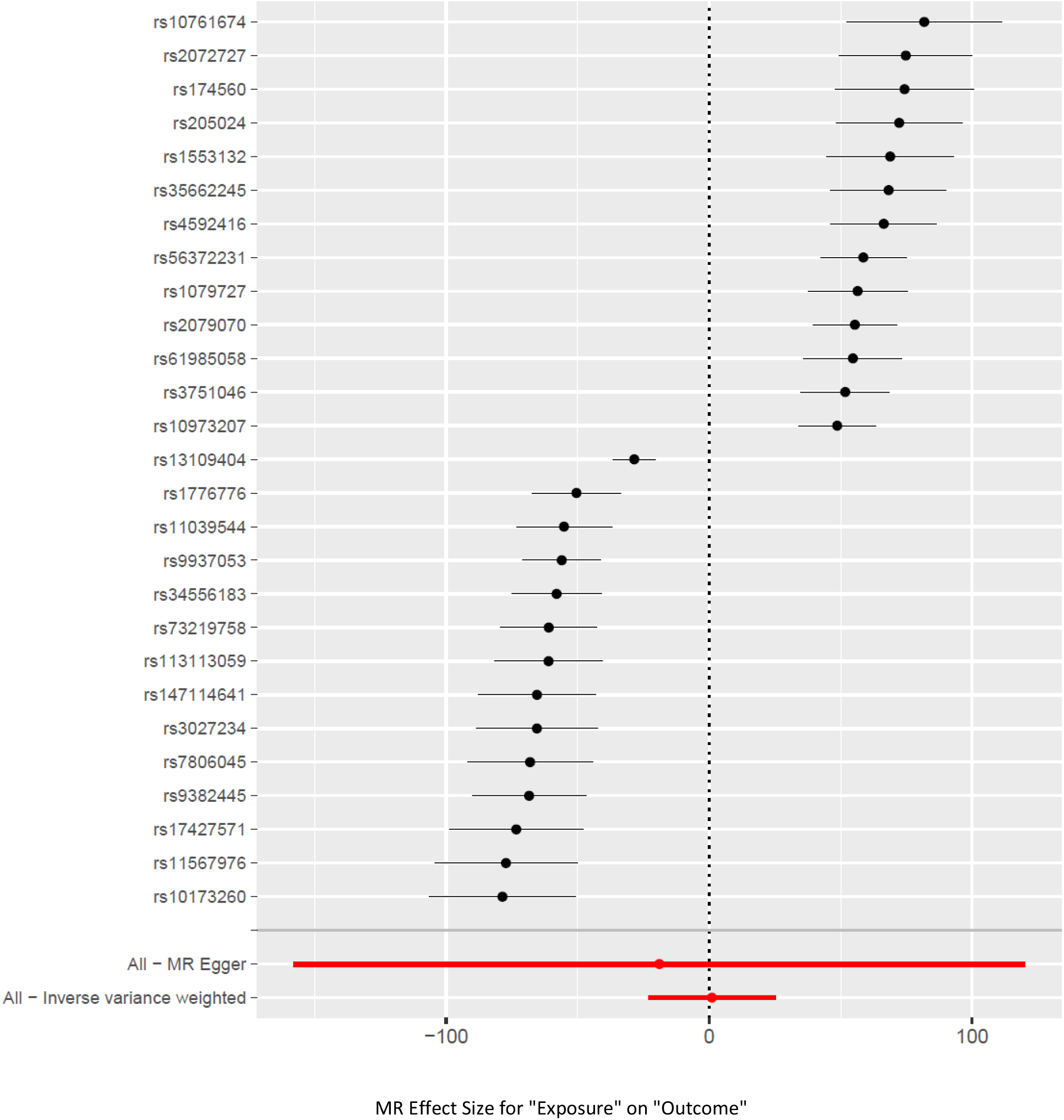
Forest Plot of the causal effect of sleep duration (exposure) on AUD (outcome)

There was a nominally significant negative causal effect of insomnia on AUDIT-C score (16 SNPs; beta = -0.45; SE = 0.17; p = 0.008; Supplementary Figure 6). Sensitivity analysis using the Egger regression estimate, weighted median, simple mode, and weighted mode showed no association. Neither heterogeneity (p = 0.25) nor a horizontal pleiotropic effect (p < 0.80) was seen, Supplementary Table 5. There was no causal effect of AUDIT-C on insomnia (12 SNPs; p = 0.92; Supplementary Table 6).

**Figure 6.**
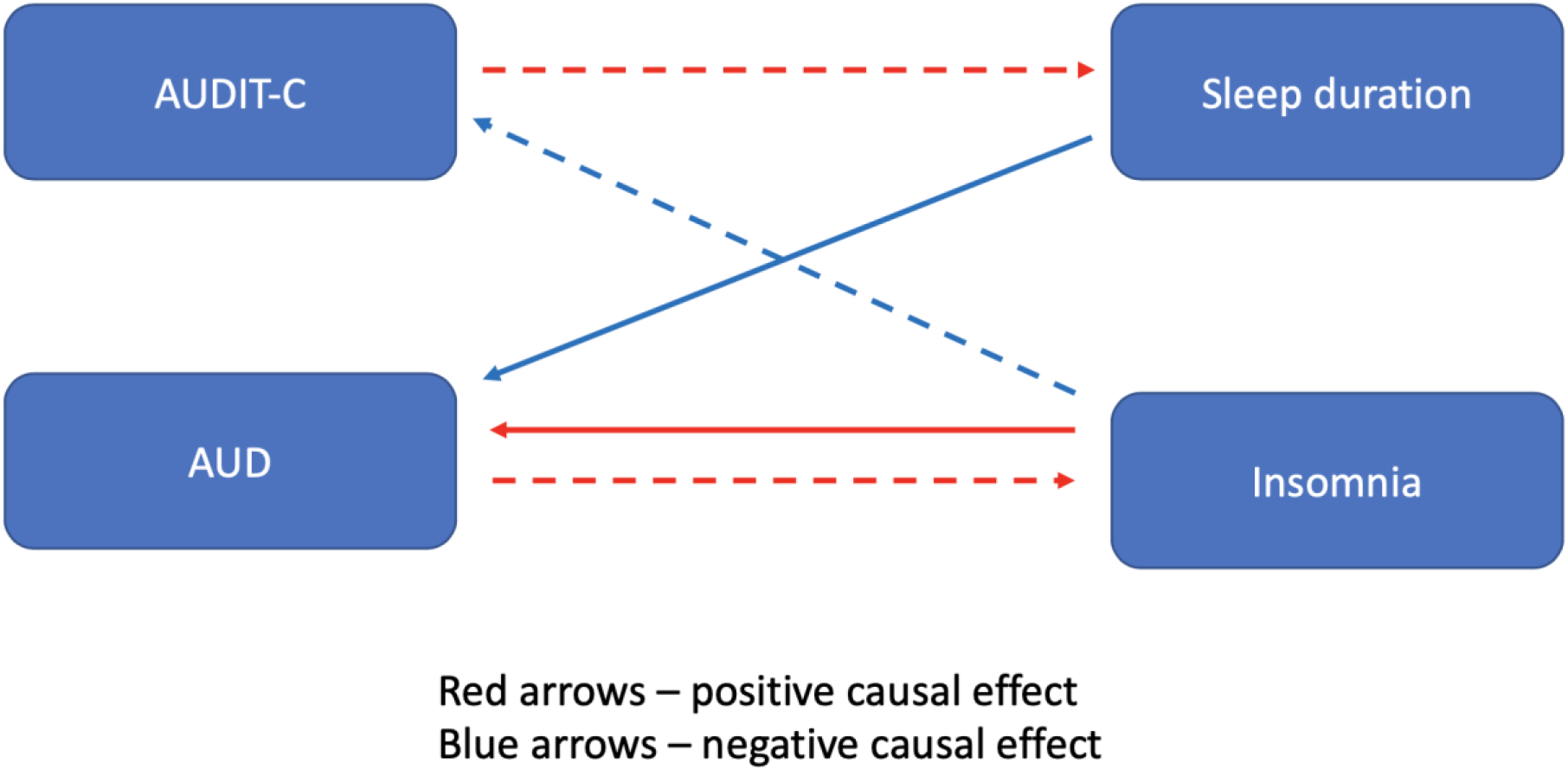
A conceptual diagram of the relationships between sleep (insomnia and sleep duration) and alcohol (AUD and AUDIT-C) variables Footnote: AUD = Alcohol Use Disorder; AUDIT-C = Alcohol Use Disorder Identification Test – Consumption. Solid arrows depict statistically significant associations. Broken arrows depict nominally significant associations.

### Sleep Duration

#### Sleep Duration PRS is not associated with AUD but is associated with alcohol consumption

The PRS for sleep duration did not show an overall significant association with AUD (Bonferroni-adjusted p-value of 0.0056), though there was a non-significant trend for a negative association with AUD at a p-value cutoff of 0.001 (OR = 0.98, p = 0.006; Figure 3c and Supplementary Table 7). The PRS for sleep duration was positively associated with AUDIT-C at two significance thresholds, the most significant of which was p = 1 × 10^−6^ (Beta = 0.01, p = 0.0009; Figure 3d and Supplementary Table 7).

#### Mendelian randomization analyses support the causal effect of sleep duration on AUD but not the reverse

The mode-based estimator using 27 SNPs showed a significant negative causal effect of sleep duration on AUD (beta = -63.05; SE = 3.54; p = 4.55 × 10^−16^; Supplementary Table 8). The causal effect of sleep duration on AUD was significant in sensitivity analyses with the weighted mode (beta = -51.66; SE = 2.68; p = 6.21 × 10^−17^) and weighted median estimator (beta = -21.78; SE = 3.80; p = 1.02 × 10^−8^) (Supplementary Table 8, Supplementary Figure 7). In single-SNP analyses all the SNPs had a significant effect, with p-values ranging from 1.41 × 10^−8^ to 1.55 × 10^−13^. None of the 27 SNPs in the leave-one-out sensitivity analysis resulted in a significantly unbalanced model (Supplementary Figure 8). The Cochrane’s Q test (p <0.0001) showed the presence of heterogeneity, with 14/27 SNPs showing an inverse association (Figure 5) but no horizontal pleiotropy was seen (p = 0.78). However, AUD was not causally associated with sleep duration (10 SNPs, p = 0.34 (Supplementary Table 9)).

#### Other findings

AUDIT-C showed a nominally positive causal effect on sleep duration (12 SNPs, Beta = 0.10, SE = 0.04, p = 0.03 (Supplementary Table 10 and Supplementary Figures 9 and 10), although heterogeneity was seen (Cochrane’s Q-test p= 1.13 × 10^−21^), but no horizontal pleiotropic effect (p = 0.49). Sensitivity analyses showed no association and there was heterogeneity in the data. Sleep duration was not significantly causally associated with AUDIT-C (Supplementary Table 11).

## Discussion

Using data from four of the largest available GWASs of insomnia, sleep duration, AUD, and AUDIT-C scores in EA individuals, we evaluated whether genetic risk for sleep-related traits was associated with alcohol-related traits. We also investigated causal relationships between the sleep- and alcohol-related variables using bi-directional Mendelian Randomization studies. We found that higher insomnia PRS was associated with greater risk of AUD and lower AUDIT-C score. MR studies showed a significant positive causal effect of insomnia on AUD, though the relationship was mitigated by heterogeneity. AUD also showed a small but statistically reliable positive causal effect on insomnia. The sleep duration PRS was positively associated with AUDIT-C. There was a nominally positive causal effect of AUDIT-C on sleep duration, but no causal effect of sleep duration on AUDIT-C. Finally, although shorter sleep duration had a positive causal effect on AUD, the reverse effect was not present.

Our PRS analysis demonstrated that genetic risk for sleep duration abnormalities is associated with increased risk of alcohol consumption. We also found that sleep duration has a negative causal effect on AUD, i.e., individuals with insufficient sleep duration are at increased risk of developing AUD. Thus, individuals at high genetic risk of decreased sleep duration are at increased risk of heavier alcohol consumption, which could lead to AUD, as also seen from the findings in the MR analysis.^38,39^ Alcohol consumption did not show a robust causal association with sleep duration in the MR investigation. These results are consistent with prior epidemiologic literature that shows that a habitual average sleep duration of less than 6 hours a night is associated with increased risk for greater alcohol consumption, cardiometabolic and neuropsychiatric disorders, and mortality,.^40^ Further, individuals with AUD or those recovering from AUD were 1.3-1.9 times more likely to report short sleep duration (< 6 hours a night) than those without AUD. These risk estimates increased to 2.6-2.9 times for individuals sleeping less than 5 hours.^13^

Epidemiological studies have also shown that insomnia is a risk factor for the development of AUD.^38,39^ This association may result from a greater preference for alcohol,^41-44^ a more rapid development of tolerance to the hypnotic effect of alcohol,^45^ and an escalation to heavy drinking that leads to AUD among susceptible individuals.^6^ However, the relationship between insomnia and AUD may be confounded by other co-occurring conditions, such as psychiatric symptoms.^46,47^ Similarly, a large GWAS of insomnia showed a substantial genetic overlap between insomnia and metabolic traits, such as type 2 diabetes, waist-hip ratio, and body mass index and strong genetic correlations with depressive symptoms, anxiety disorder, and subjective well-being.^20^ These co-occurring conditions may explain the PRS findings reported here that the genetic risk for insomnia is associated with AUD and is consistent with findings in prior clinical studies.^7^ The heterogeneity in the MR analyses may be related to variation in the diagnosis of AUD, because the diagnoses was derived from ICD-9 or ICD-10 clinical codes from their clinical charts within the VA, rather than being obtained through administration of a structured interview or other method aimed at ensuring diagnostic reliability.

AUDIT-C scores were positively associated with the sleep duration PRS and negatively associated with insomnia PRS, contrary to the existing clinical literature showing an inverse association between alcohol consumption and sleep duration, but a positive association between drinking and insomnia. ^6,11-14,48^ AUDIT-C data for this study were extracted from clinical charts of Veterans administered as a part of their annual routine screening for alcohol consumption. The heterogeneous nature of AUDIT-C reports, as seen by the lack of a dose-response curve of the AUDIT-C scale scores with the insomnia PRS in Supplementary Figure 1, and the higher prevalence of insomnia in Veterans relative to individuals in the community ^49,50^ may have introduced a bias in the relationship between AUDIT-C and insomnia in this dataset. The weak pleiotropic effects between sleep duration and alcohol-related measures may also reflect a greater impact of environmental factors than genetic factors as being responsible for these associations.

In contrast to prior findings of insomnia showing a clear causal effect on AUD,^27^ we found the relationship to be heterogeneous. Our study used summary statistics from a larger sample of individuals (202,004 compared to 46,568 in the prior study^27^). Further, the AUD diagnoses in the MVP GWAS were clinically based and required the presence of either one instance of an inpatient diagnosis or two outpatient diagnoses made at different times. In contrast, the prior study^27^ used GWAS data from participants whose AUD diagnoses were made using clinician ratings or semi-structured interviews.^51^ Finally, the earlier study may have introduced confounding by using the Steiger filtering methodology in their MR analysis.^27^ In Steiger filtering, SNPs that explain more variance in the outcome than the exposure are excluded. In contrast, in the present analysis, SNPs that explained significantly more variance in the exposure (p<0.05) were retained in our MR methodology.

Our study has limitations. Although we included sensitivity analyses to assess horizontal pleiotropy, we could not exclude this potential bias completely, using mode-based estimators as needed. We also used a subjective measure of sleep duration, which in prior research has only a small-to-moderate correlation with measured sleep duration.^52,53^ In contrast to subjective sleep duration, the insomnia question in the UK Biobank has 98% sensitivity and 96% specificity in discriminating patients with insomnia disorder from unaffected controls.^54^ Because we conducted our analyses using data from the UK Biobank and MVP the results are not representative of the general population because of selection biases in the samples. This is evidenced by the fact that US Veterans have a higher prevalence of insomnia, heavy drinking, and AUD than the general population. ^49,50,55-59^ Future studies should evaluate these relationships in ancestral groups other than European. Nonetheless, this is the first study to evaluate the association between PRS based on common sleep-related traits and alcohol-related traits. It is also the first to assess causal relationships between subjective sleep duration and alcohol-related traits.

## Conclusion

The genetic determinants of sleep traits and alcohol-related traits are partially overlapping. The genetic risk for insomnia has pleiotropic effects on alcohol consumption and AUD, and insomnia has a causal effect on AUD. Short sleep duration may causally influence AUD risk and interventions that increase sleep duration may be a promising therapeutic target for AUD.

## Data Availability

All data produced in the present work are contained in the manuscript.

https://www.ebi.ac.uk/gwas/home

## Acknowledgment

This work was supported by the following grants: 1IK2CX000855 (S.C.); 1 I01 CX001957 (S.C.); I01 BX003341 (H.R.K); R01 HL143790 (P.R.G), R01 NR018836 (P.R.G.); 1K01 AA028292 (R.L.K.); and the Veterans Integrated Service Network 4 Mental Illness Research, Education, and Clinical Center. The content of this publication does not represent the views of the Department of Veterans Affairs, the US government, Perelman School of Medicine, or other participating institutions.

## References

1. Boersma P, Villarroel MA, Vahratian A. Heavy Drinking Among U.S. Adults, 2018. Centers for Disease Control and Prevention. https://www.cdc.gov/nchs/products/databriefs/db374.htm. Published 2020. Accessed April 26, 2021.

2. Grant BF, Chou SP, Saha TD, et al. Prevalence of 12-Month Alcohol Use, High-Risk Drinking, and DSM-IV Alcohol Use Disorder in the United States, 2001-2002 to 2012-2013: Results From the National Epidemiologic Survey on Alcohol and Related Conditions. JAMA Psychiatry. 2017;74(9):911–923.

3. Gordon AJ, Fiellin DA, Friedmann PD, et al. Update in addiction medicine for the primary care clinician. J Gen Intern Med. 2008;23(12):2112–2116.

4. N.I.A.A.A. Rethinking Drinking. https://www.rethinkingdrinking.niaaa.nih.gov/How-much-is-too-much/Whats-the-harm/What-Are-The-Risks.aspx. Accessed April 24, 2021.

5. Dawson DA, Li TK, Grant BF. A prospective study of risk drinking: at risk for what? Drug Alcohol Depend. 2008;95(1-2):62–72.

6. Haario P, Rahkonen O, Laaksonen M, Lahelma E, Lallukka T. Bidirectional associations between insomnia symptoms and unhealthy behaviours. Journal of sleep research. 2013;22(1):89–95.

7. Chakravorty S, Chaudhary NS, Brower KJ. Alcohol Dependence and Its Relationship With Insomnia and Other Sleep Disorders. Alcohol Clin Exp Res. 2016.

8. Kline C. Sleep Duration. In: Gellman MD, Turner JR, eds. Encyclopedia of Behavioral Medicine. New York, NY: Springer; 2013:1808–1810.

9. Consensus Conference P, Watson NF, Badr MS, et al. Joint Consensus Statement of the American Academy of Sleep Medicine and Sleep Research Society on the Recommended Amount of Sleep for a Healthy Adult: Methodology and Discussion. J Clin Sleep Med. 2015;11(8):931–952.

10. Consensus Conference P, Watson NF, Badr MS, et al. Recommended Amount of Sleep for a Healthy Adult: A Joint Consensus Statement of the American Academy of Sleep Medicine and Sleep Research Society. J Clin Sleep Med. 2015;11(6):591–592.

11. Krueger PM, Friedman EM. Sleep duration in the United States: a cross-sectional population-based study. Am J Epidemiol. 2009;169(9):1052–1063.

12. Palmer CD, Harrison GA, Hiorns RW. Association between smoking and drinking and sleep duration. Annals of human biology. 1980;7(2):103–107.

13. John U, Meyer C, Rumpf HJ, Hapke U. Relationships of psychiatric disorders with sleep duration in an adult general population sample. J Psychiatr Res. 2005;39(6):577–583.

14. Schuckit MA, Bernstein LI. Sleep time and drinking history: a hypothesis. The American journal of psychiatry. 1981;138(4):528–530.

15. Chaput JP, McNeil J, Despres JP, Bouchard C, Tremblay A. Short sleep duration is associated with greater alcohol consumption in adults. Appetite. 2012;59(3):650–655.

16. Wray N, Visscher P. Estimating trait heritability.. Nature Education. 2008;1(1):29.

17. Kranzler HR, Zhou H, Kember RL, et al. Genome-wide association study of alcohol consumption and use disorder in 274,424 individuals from multiple populations. Nat Commun. 2019;10(1):1499.

18. Sanchez-Roige S, Palmer AA, Fontanillas P, et al. Genome-Wide Association Study Meta-Analysis of the Alcohol Use Disorders Identification Test (AUDIT) in Two Population-Based Cohorts. Am J Psychiatry. 2019;176(2):107–118.

19. Vrieze SI, McGue M, Miller MB, Hicks BM, Iacono WG. Three mutually informative ways to understand the genetic relationships among behavioral disinhibition, alcohol use, drug use, nicotine use/dependence, and their co-occurrence: twin biometry, GCTA, and genome-wide scoring. Behav Genet. 2013;43(2):97–107.

20. Jansen PR, Watanabe K, Stringer S, et al. Genome-wide analysis of insomnia in 1,331,010 individuals identifies new risk loci and functional pathways. Nat Genet. 2019;51(3):394–403.

21. Lane JM, Jones SE, Dashti HS, et al. Biological and clinical insights from genetics of insomnia symptoms. Nat Genet. 2019;51(3):387–393.

22. Dashti HS, Jones SE, Wood AR, et al. Genome-wide association study identifies genetic loci for self-reported habitual sleep duration supported by accelerometer-derived estimates. Nat Commun. 2019;10(1):1100.

23. Choi SW, Mak TS, O’Reilly PF. Tutorial: a guide to performing polygenic risk score analyses. Nat Protoc. 2020;15(9):2759–2772.

24. Davies NM, Holmes MV, Davey Smith G. Reading Mendelian randomisation studies: a guide, glossary, and checklist for clinicians. BMJ. 2018;362:k601.

25. Hemani G, Zheng J, Elsworth B, et al. The MR-Base platform supports systematic causal inference across the human phenome. Elife. 2018;7.

26. Davey Smith G, Hemani G. Mendelian randomization: genetic anchors for causal inference in epidemiological studies. Hum Mol Genet. 2014;23(R1):R89–98.

27. Pasman JA, Smit DJA, Kingma L, Vink JM, Treur JL, Verweij KJH. Causal relationships between substance use and insomnia. Drug Alcohol Depend. 2020;214:108151.

28. Torkamani A, Wineinger NE, Topol EJ. The personal and clinical utility of polygenic risk scores. Nat Rev Genet. 2018;19(9):581–590.

29. Sudlow C, Gallacher J, Allen N, et al. UK biobank: an open access resource for identifying the causes of a wide range of complex diseases of middle and old age. PLoS Med. 2015;12(3):e1001779.

30. Gaziano JM, Concato J, Brophy M, et al. Million Veteran Program: A mega-biobank to study genetic influences on health and disease. J Clin Epidemiol. 2016;70:214–223.

31. Bush K, Kivlahan DR, McDonell MB, Fihn SD, Bradley KA. The AUDIT alcohol consumption questions (AUDIT-C): an effective brief screening test for problem drinking. Ambulatory Care Quality Improvement Project (ACQUIP). Alcohol Use Disorders Identification Test. Arch Intern Med. 1998;158(16):1789–1795.

32. Bulik-Sullivan B, Finucane HK, Anttila V, et al. An atlas of genetic correlations across human diseases and traits. Nat Genet. 2015;47(11):1236–1241.

33. Yavorska OO, Burgess S. MendelianRandomization: an R package for performing Mendelian randomization analyses using summarized data. Int J Epidemiol. 2017;46(6):1734–1739.

34. Burgess S, Labrecque JA. Mendelian randomization with a binary exposure variable: interpretation and presentation of causal estimates. Eur J Epidemiol. 2018;33(10):947–952.

35. Bowden J, Hemani G, Davey Smith G. Invited Commentary: Detecting Individual and Global Horizontal Pleiotropy in Mendelian Randomization-A Job for the Humble Heterogeneity Statistic? Am J Epidemiol. 2018;187(12):2681–2685.

36. Purcell S, Neale B, Todd-Brown K, et al. PLINK: a tool set for whole-genome association and population-based linkage analyses. Am J Hum Genet. 2007;81(3):559–575.

37. Team RC. R: A language and environment for statistical computing. 2018.

38. Ford DE, Kamerow DB. Epidemiologic study of sleep disturbances and psychiatric disorders. An opportunity for prevention? JAMA. 1989;262(11):1479–1484.

39. Weissman MM, Greenwald S, Nino-Murcia G, Dement WC. The morbidity of insomnia uncomplicated by psychiatric disorders. Gen Hosp Psychiatry. 1997;19(4):245–250.

40. Grandner MA, Patel NP, Gehrman PR, Perlis ML, Pack AI. Problems associated with short sleep: bridging the gap between laboratory and epidemiological studies. Sleep medicine reviews. 2010;14(4):239–247.

41. Roehrs T, Papineau K, Rosenthal L, Roth T. Ethanol as a hypnotic in insomniacs: self administration and effects on sleep and mood. Neuropsychopharmacology. 1999;20(3):279–286.

42. Ancoli-Israel S, Roth T. Characteristics of insomnia in the United States: results of the 1991 National Sleep Foundation Survey. I. Sleep. 1999;22 Suppl 2:S347–353.

43. Johnson EO, Roehrs T, Roth T, Breslau N. Epidemiology of alcohol and medication as aids to sleep in early adulthood. Sleep: Journal of Sleep Research & Sleep Medicine. 1998;21(2):178–186.

44. Kaneita Y, Uchiyama M, Takemura S, et al. Use of alcohol and hypnotic medication as aids to sleep among the Japanese general population. Sleep Medicine. 2007;8(7-8):723–732.

45. Roehrs T, Roth T. Insomnia as a path to alcoholism: tolerance development and dose escalation. Sleep. 2018;41(8).

46. Chaudhary NS, Wong MM, Kolla BP, Kampman KM, Chakravorty S. The relationship between insomnia and the intensity of drinking in treatment-seeking individuals with alcohol dependence. Drug Alcohol Depend. 2020;215:108189.

47. Kolla BP, Mansukhani MP, Biernacka J, Chakravorty S, Karpyak VM. Sleep disturbances in early alcohol recovery: Prevalence and associations with clinical characteristics and severity of alcohol consumption. Drug Alcohol Depend. 2020;206:107655.

48. Canham SL, Kaufmann CN, Mauro PM, Mojtabai R, Spira AP. Binge drinking and insomnia in middle-aged and older adults: the Health and Retirement Study. Int J Geriatr Psychiatry. 2015;30(3):284–291.

49. Klingaman EA, Brownlow JA, Boland EM, Mosti C, Gehrman PR. Prevalence, predictors and correlates of insomnia in US army soldiers. J Sleep Res. 2018;27(3):e12612.

50. Alexander M, Ray MA, Hebert JR, et al. The National Veteran Sleep Disorder Study: Descriptive Epidemiology and Secular Trends, 2000-2010. Sleep. 2016;39(7):1399–1410.

51. Walters RK, Polimanti R, Johnson EC, et al. Transancestral GWAS of alcohol dependence reveals common genetic underpinnings with psychiatric disorders. Nat Neurosci. 2018;21(12):1656–1669.

52. Lauderdale DS, Knutson KL, Yan LL, Liu K, Rathouz PJ. Self-reported and measured sleep duration: how similar are they? Epidemiology. 2008;19(6):838–845.

53. Jones SE, van Hees VT, Mazzotti DR, et al. Genetic studies of accelerometer-based sleep measures yield new insights into human sleep behaviour. Nat Commun. 2019;10(1):1585.

54. Hammerschlag AR, Stringer S, de Leeuw CA, et al. Genome-wide association analysis of insomnia complaints identifies risk genes and genetic overlap with psychiatric and metabolic traits. Nat Genet. 2017;49(11):1584–1592.

55. Pigeon WR, Campbell CE, Possemato K, Ouimette P. Longitudinal relationships of insomnia, nightmares, and PTSD severity in recent combat veterans. J Psychosom Res. 2013;75(6):546–550.

56. Mustafa M, Erokwu N, Ebose I, Strohl K. Sleep problems and the risk for sleep disorders in an outpatient veteran population. Sleep Breath. 2005;9(2):57–63.

57. Tsai J, Kasprow WJ, Rosenheck RA. Alcohol and drug use disorders among homeless veterans: prevalence and association with supported housing outcomes. Addict Behav. 2014;39(2):455–460.

58. Vazan P, Golub A, Bennett AS. Substance use and other mental health disorders among veterans returning to the inner city: prevalence, correlates, and rates of unmet treatment need. Subst Use Misuse. 2013;48(10):880–893.

59. Grant BF, Goldstein RB, Saha TD, et al. Epidemiology of DSM-5 Alcohol Use Disorder: Results From the National Epidemiologic Survey on Alcohol and Related Conditions III. JAMA Psychiatry. 2015;72(8):757–766.

