## Supplementary Figures for "The Relationship between Alcohol- and Sleep-related Traits: Results from Polygenic Risk Score Analyses and Mendelian Randomization Studies"

Supplementary Figure 1. The association between insomnia Polygenic Risk Score and Alcohol Use Disorders Identification Test-Consumption (AUDIT-C) score categories

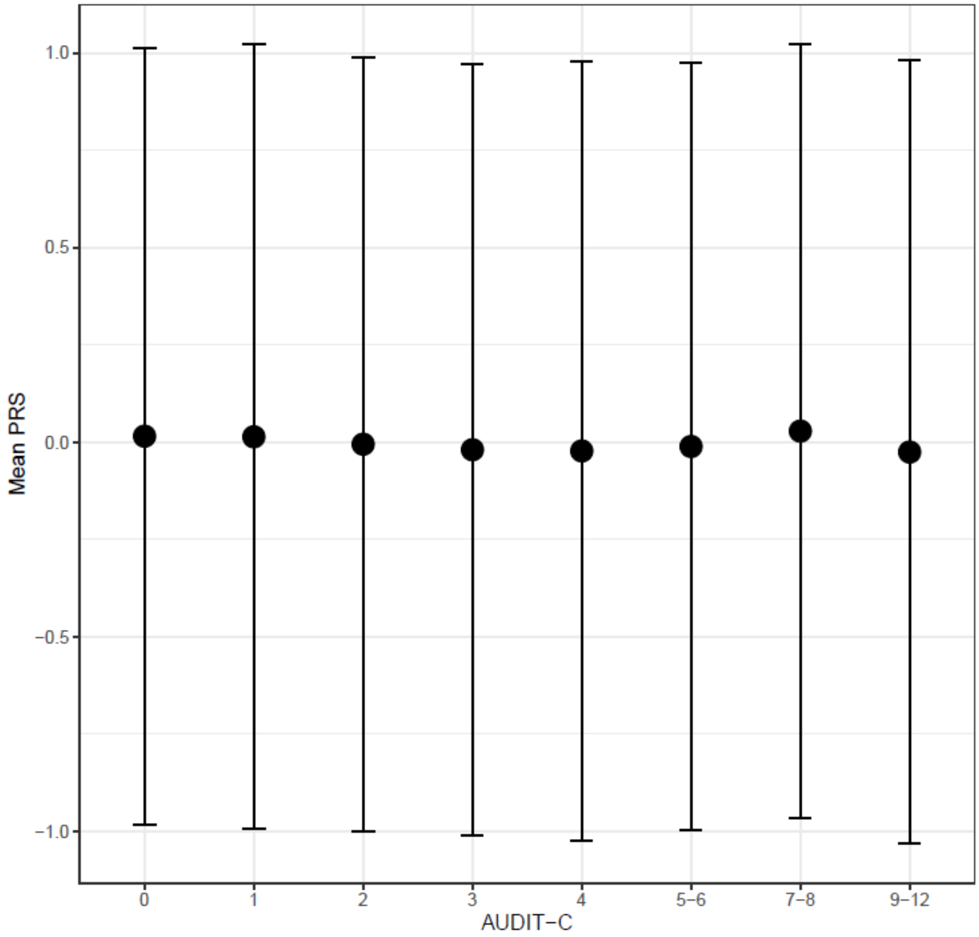

Supplementary Figure 2. Scatter plot of Insomnia (exposure) on Alcohol Use Disorder (AUD, outcome)

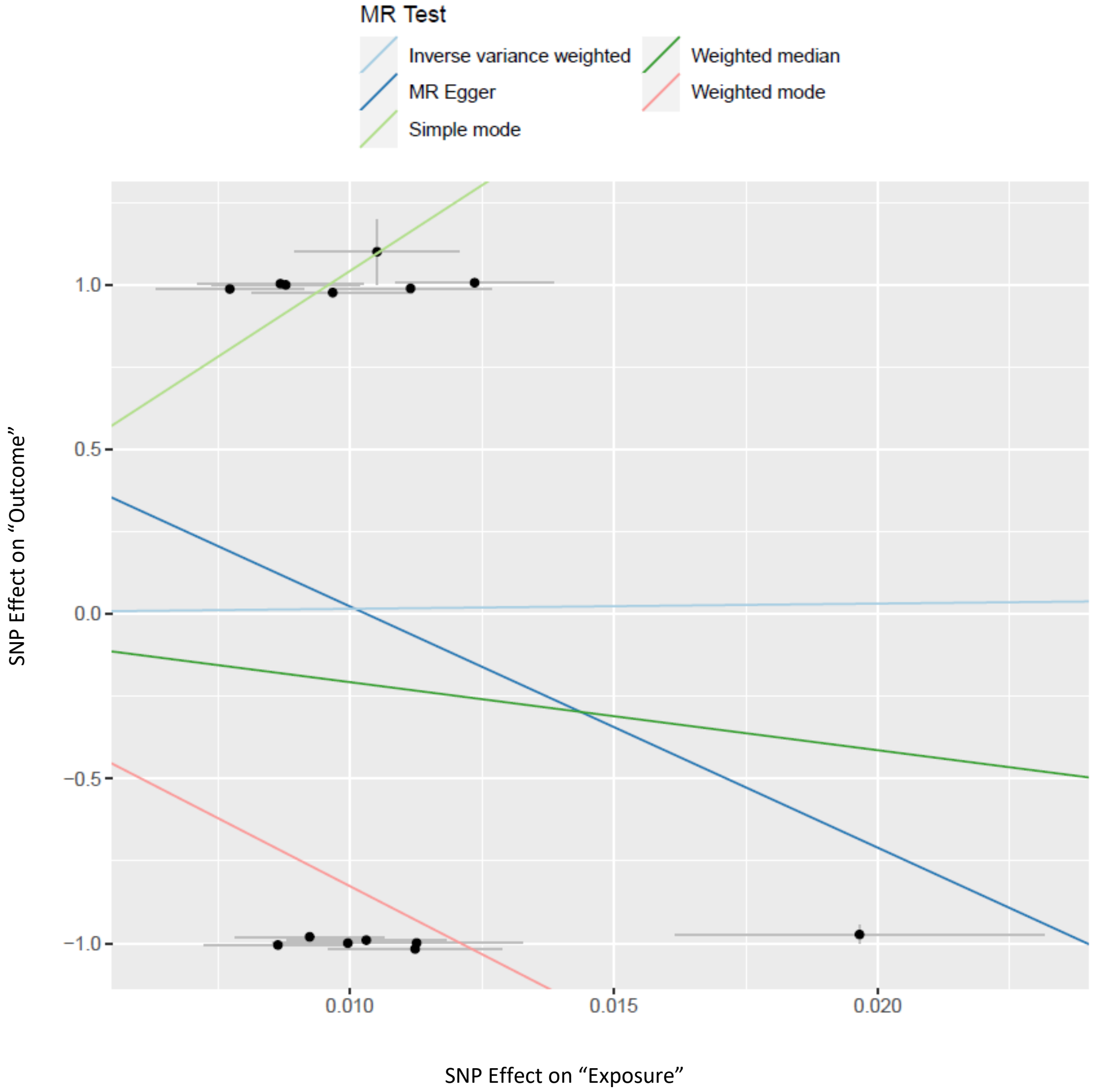

Supplementary Figure 3. Leave One Out sensitivity analysis involving Insomnia (exposure) on Alcohol Use Disorder (AUD, outcome)

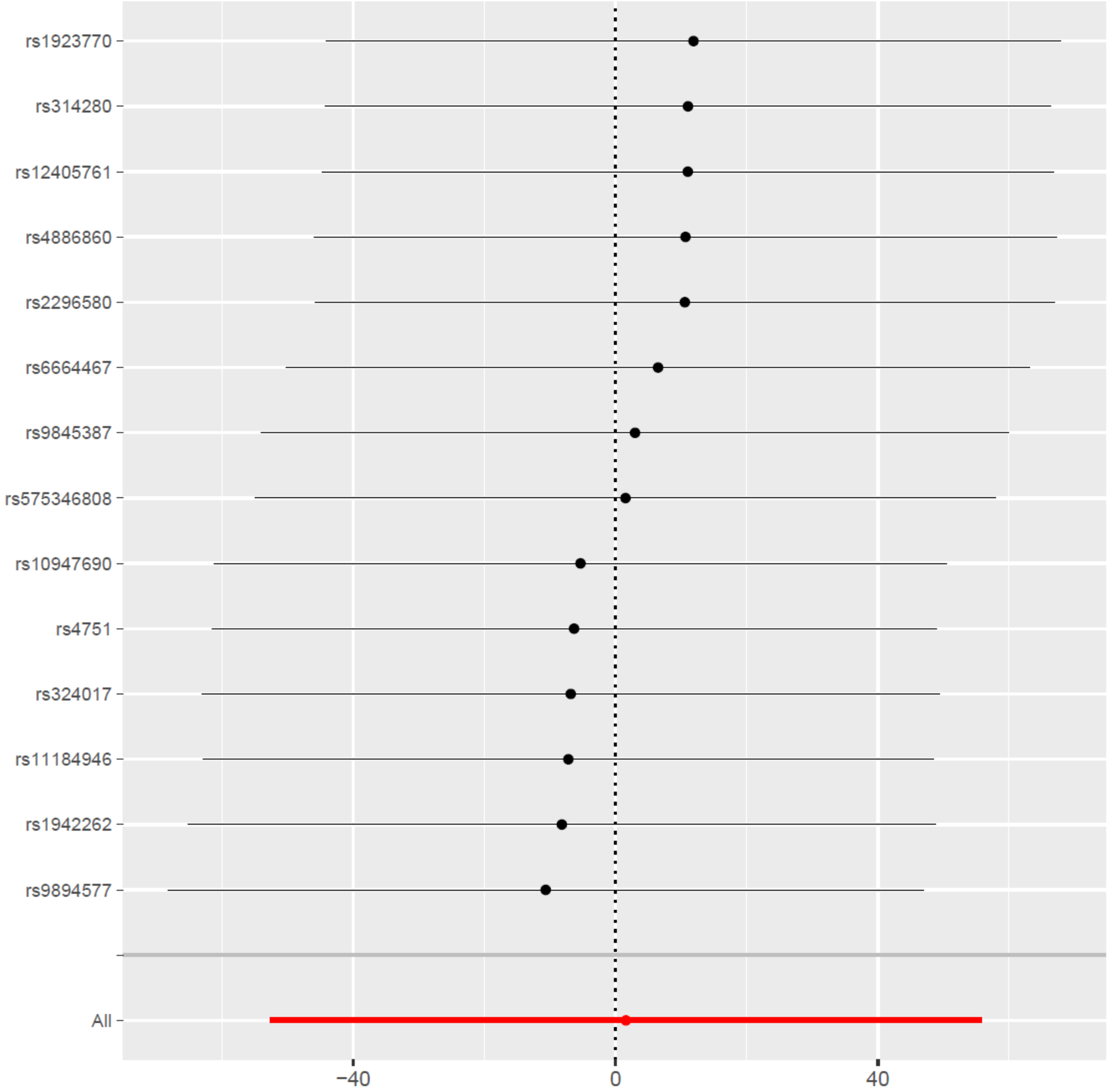

Supplementary Figure 4. Forest plot of Alcohol Use Disorder (AUD, exposure) on Insomnia (outcome)

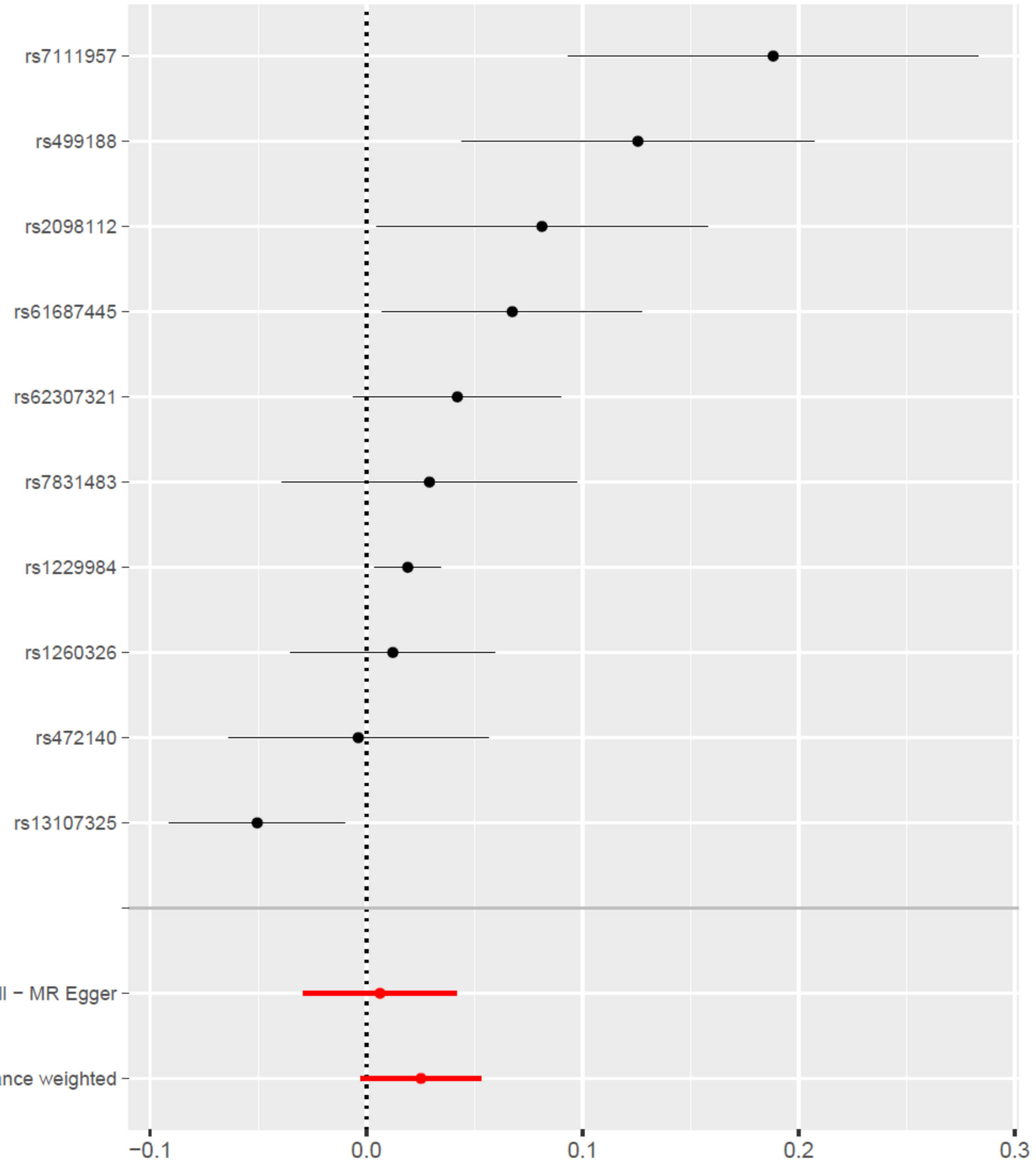

Supplementary Figure 5. Scatter plot of Alcohol Use Disorder (AUD, exposure) on Insomnia (outcome)

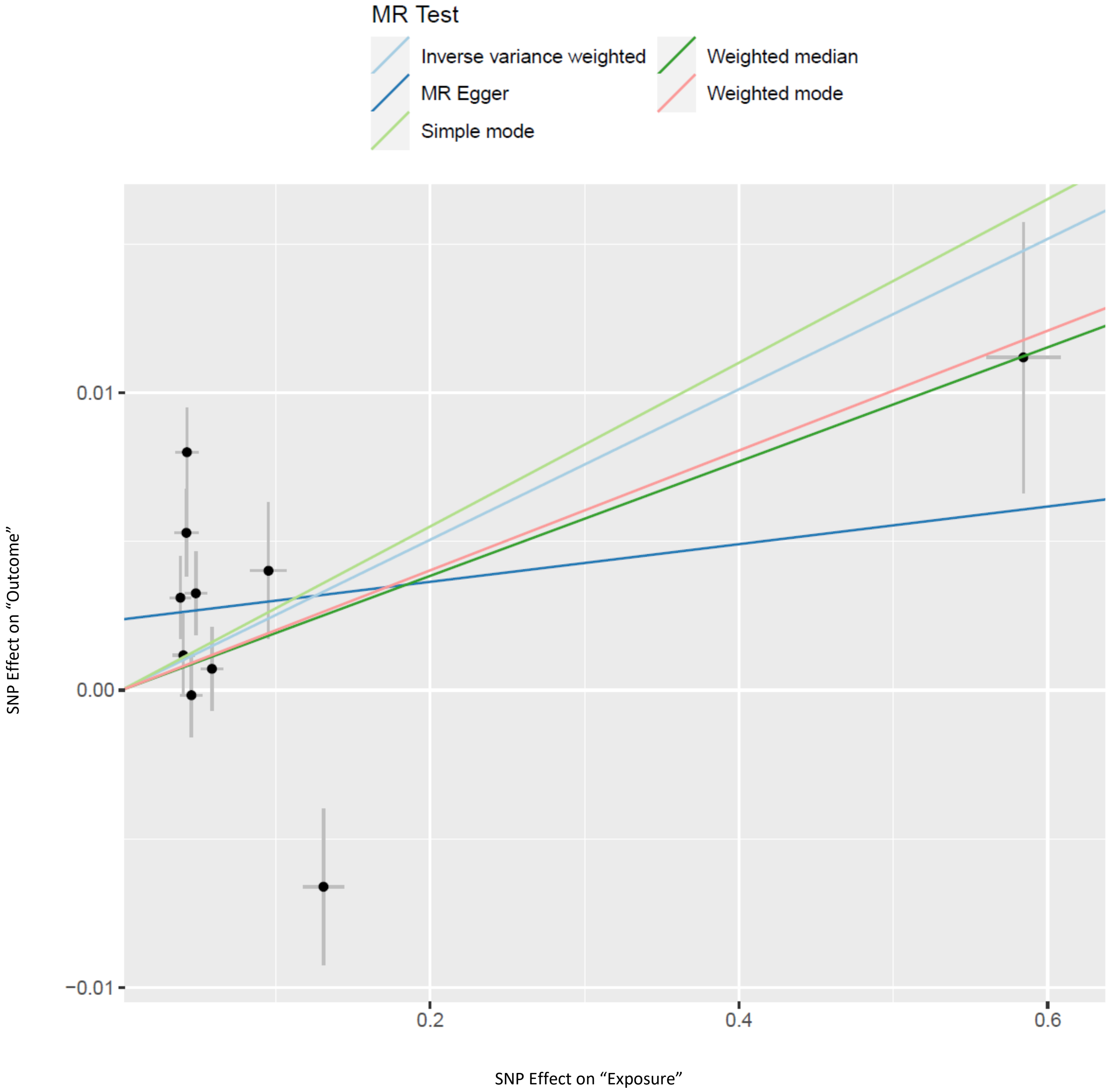

Supplementary Figure 6. Forest Plot of the causal effect of Insomnia (exposure) on drinking (AUDIT-C, outcome)

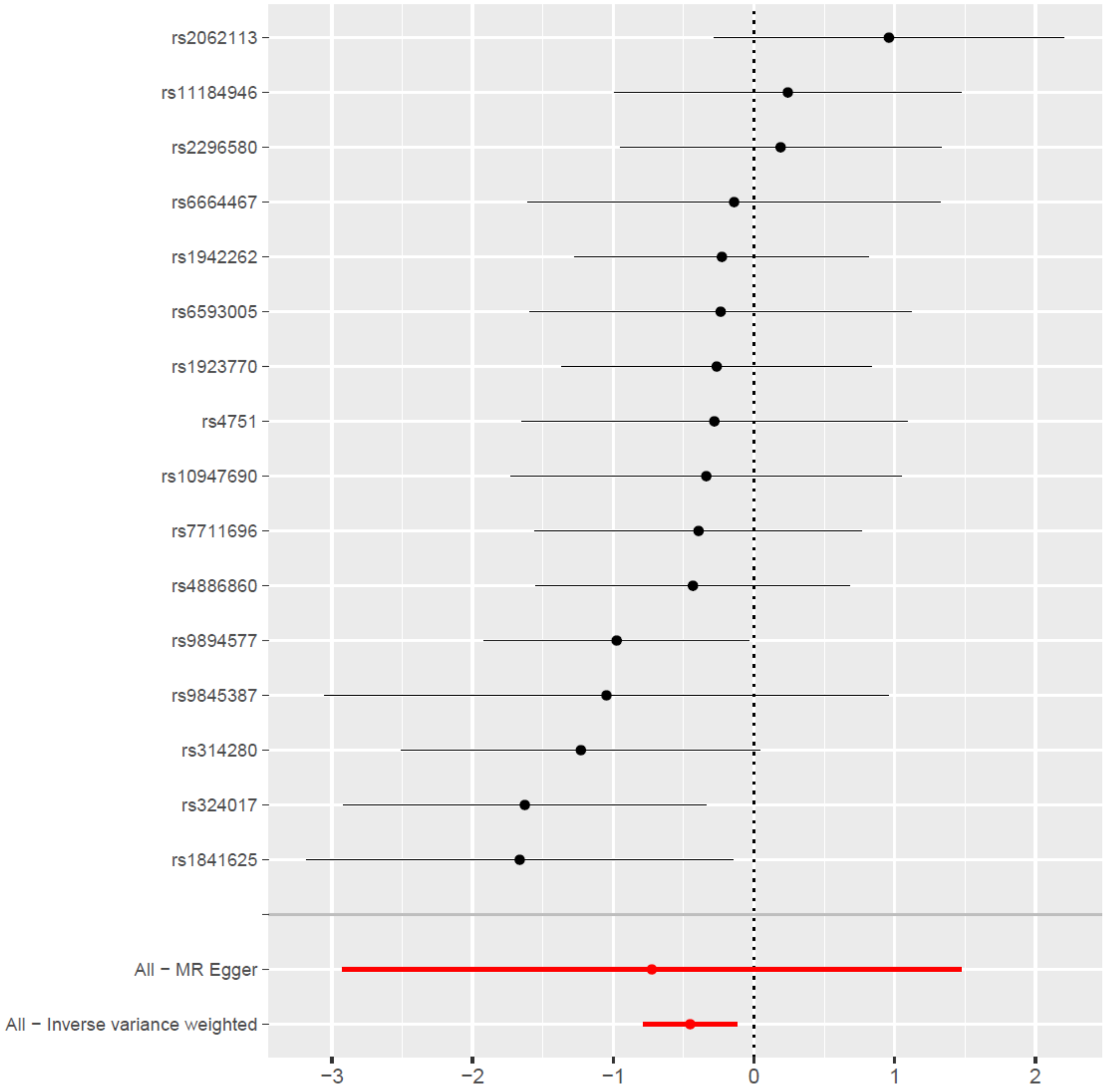

Supplementary Figure 7. Scatter plot of sleep duration (exposure) on Alcohol Use Disorder (AUD, outcome)

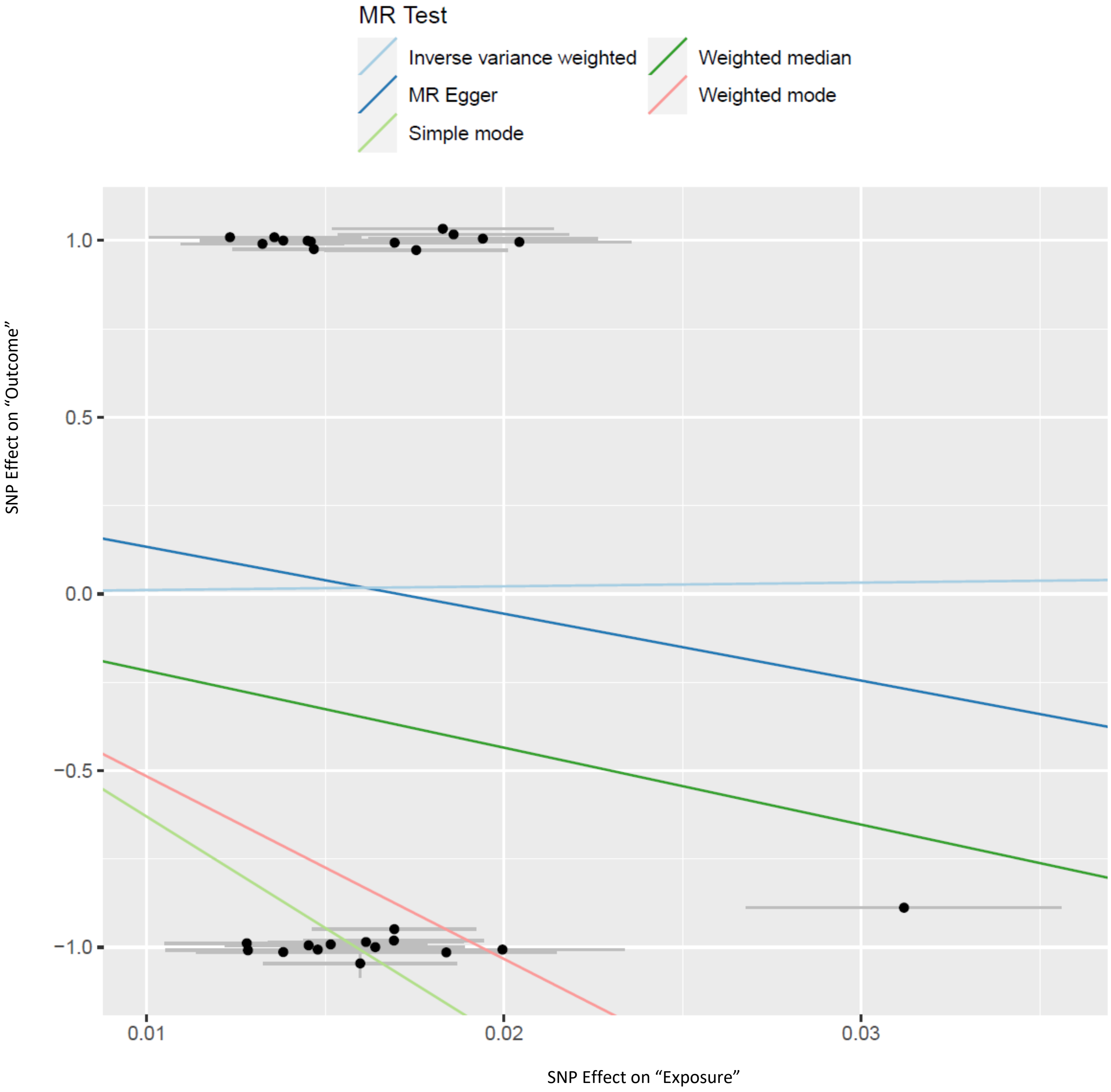

Supplementary Figure 8. Leave One Out sensitivity analysis of sleep duration (exposure) on Alcohol Use Disorder (AUD, outcome)

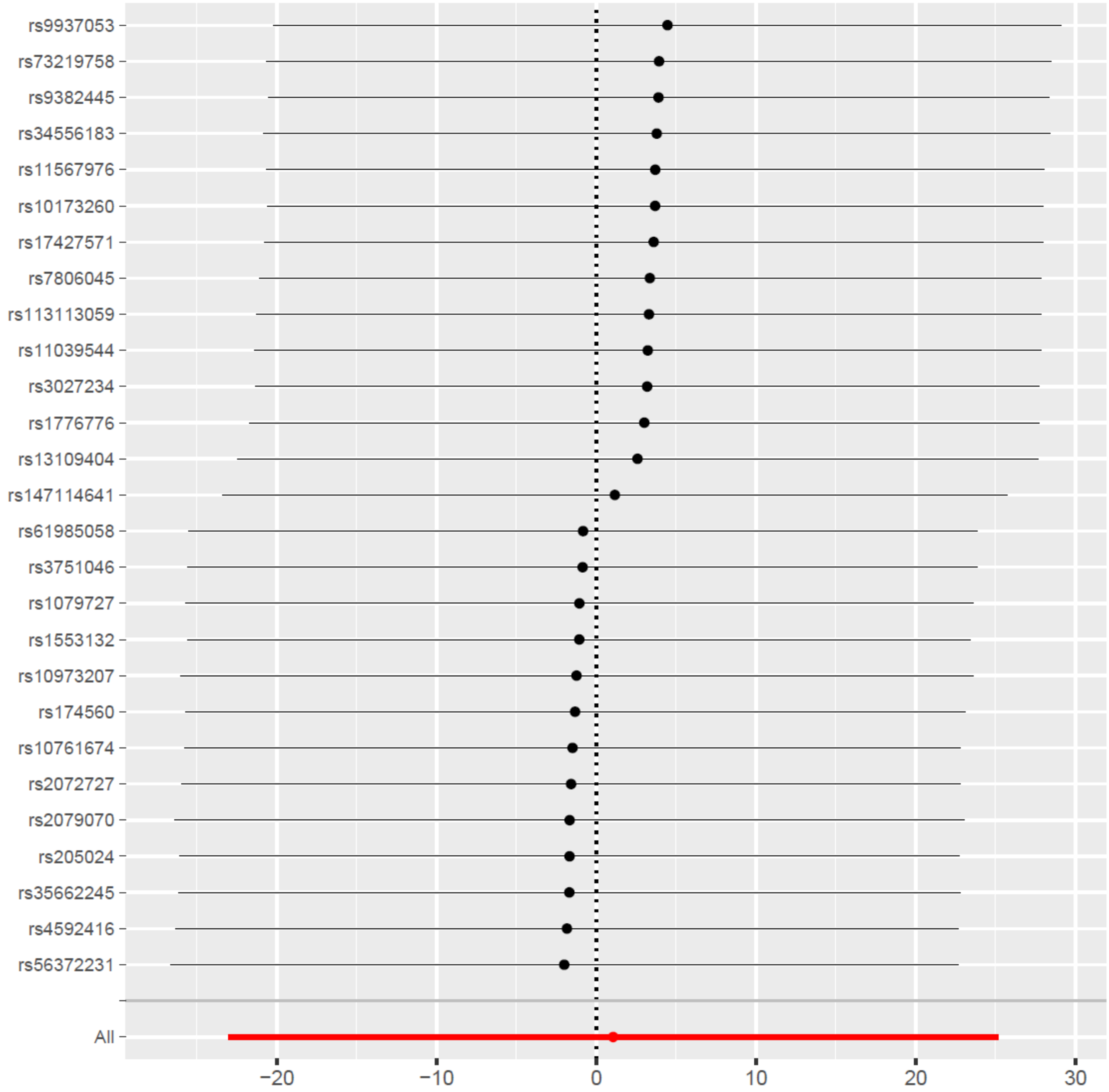

Supplementary Figure 9. Forest plot Alcohol Use Disorders Identification Test-Consumption (AUDIT-C) measure (exposure) on sleep duration (outcome)

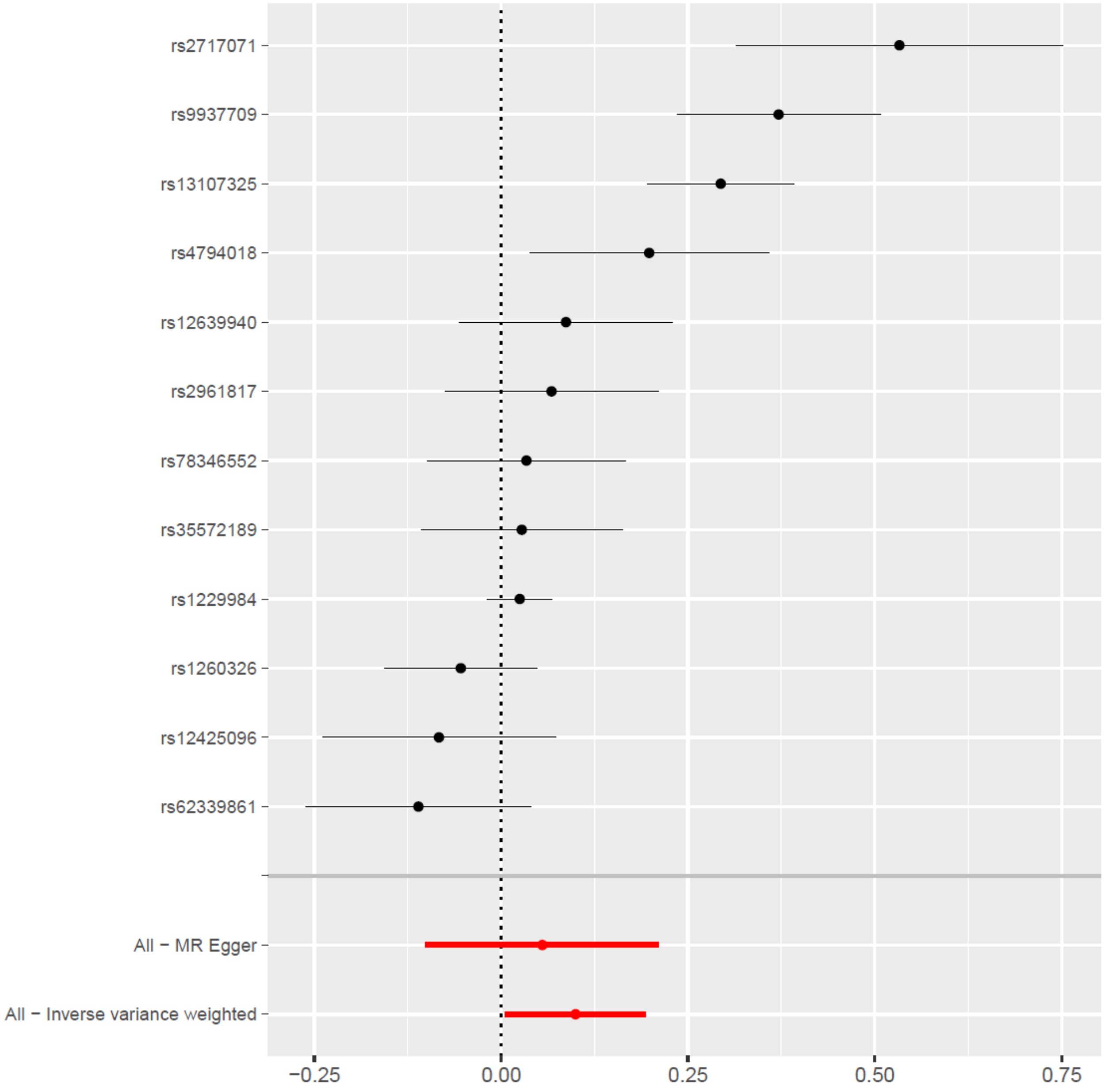

Supplementary Figure 10. Scatter plot of Alcohol Use Disorders Identification Test-Consumption (AUDIT-C) measure (exposure) on Sleep duration (outcome)

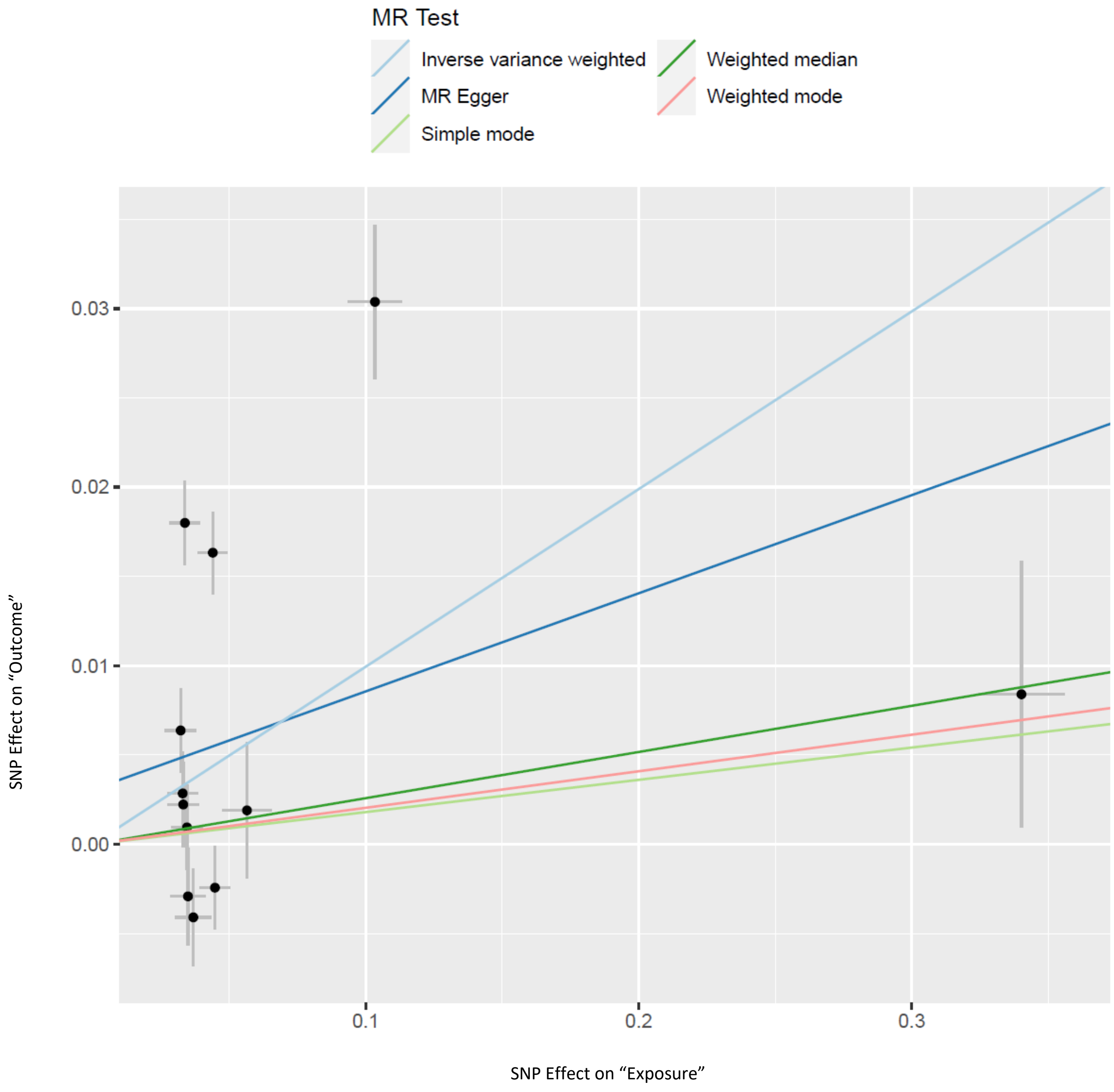
